# High prevalence of hepatitis B and HIV among women survivors of sexual violence in South Kivu province, eastern Democratic Republic of Congo

**DOI:** 10.1101/2023.09.22.23295978

**Authors:** Parvine Basimane Bisimwa, Dieudonné Bihehe Masemo, Aline Kusinza Byabene, Georges Kikuni Besulani, Théophile Kashosi Mitima, Cadeau Mugisho Matabishi, Bienfait Mitima Misuka, Omari Mukanga, Jean Paulin Mukonkole, Jean Bisimwa Nachega, Denis Mukwege, Komas Narcisse Patrice Joseph

## Abstract

**Introduction:** Limited data are available on the prevalence rates of hepatitis B and acquired immunodeficiency syndrome (AIDS) among women survivors of sexual violence (WSSV) in South Kivu province, in the eastern part of the Democratic Republic of Congo (DRC), where armed conflicts persist. Here, we aimed to assess the prevalence of these two diseases in this vulnerable local population.

**Methods:** A total of 1002 WSSV, aged from 18 to 70 years old were enrolled from May 2018 to May 2020 at three one-stop centers, set up at the Panzi, Mulamba and Bulenga hospitals. Blood samples were collected and tested for hepatitis B virus (HBV) and human immunodeficiency virus (HIV) antigens and antibodies using enzyme-linked immunoassay (ELISA) methods. Viral load quantification for HBV and HIV were performed using the GeneXpert.

**Results:** For HBV, overall prevalence was 8.9% (95% CI [7.2–10.8%]), 32.1% [29.3–35.0%], and 14.5% [12.3–16.8%] for HBsAg, anti-HBc and anti-HBs antibodies, respectively. Among the 89 HBsAg-positive patients, 17 (19.1%) were HBeAg-positive. The mean age was 40.57±14.99 years in the HBsAg-positive group (*p*=0.025). Risk factors for HBV infection were age (≥35 years) (AOR=1.83 [1.02-3.32]; *p*=0.041), having no schooling (AOR=4.14 [1.35-12.62]; *p*=0.012) or only primary school-level (AOR=4.88 [1.61-14.75]; *p*=0.005), and multiple aggressors (AOR=1.76 [1.09-2.84], *p*=0.019). The prevalence of HIV was 4.3% [3.1–5.7%]. HIV/HBV co-infection occurred only in 5 individuals (0.5%). The HBV viral load was detectable (>2,000 copies/mL) in 61.8% of HBsAg-positive subjects and 64.8% HIV-positive subjects had a high viral load (>1,000 copies/mL).

**Conclusion:** This study revealed a high prevalence of HBV and HIV infections among WSSV in South Kivu. These results highlight the urgent need for systematic screening of HBV and HIV by integrating fourth-generation ELISA tests in HIV and HBV control programs.

## Introduction

Sexually transmitted infections (STIs) constitute a heavy public health burden in many developing countries, including the Democratic Republic of Congo (DRC). Two STIs, i.e. those caused by hepatitis B virus (HBV) and human immunodeficiency virus (HIV), affect approximately 2 billion and 37.7 million people worldwide, respectively [1,2]. HBV is one of the leading infectious killers globally, with 887,000 deaths due to HBV-associated hepatic sequelae such as acute hepatitis, liver cirrhosis, and liver cancer or hepatocellular carcinoma (HCC), recorded worldwide in 2015 [1]. Its prevalence is highest in the Western Pacific and African regions, where respectively 6.2% and 6.1% of the adult population is infected [3]. The World Health Organization (WHO) has classified the DRC as an intermediate endemicity area, with 5 to 7.99% of HBV infection [1,3]. In endemicity areas, HBV infection is usually transmitted from mother to child at delivery or by horizontal transmission among children, especially during the first five years [3–5]. It is also transmitted by sexual intercourse through vaginal and seminal secretions, by tattooing and piercing with non-sterilized equipment, by blood exposure accidents, and by other infected body fluids such as saliva [1,3,4].

The Joint United Nations Program on HIV/AIDS (UNAIDS) and WHO reports show that among the 37.7 million people living with HIV worldwide, adults (36.0 million) predominate. Of these infected adults, women, and girls (53%) were the most infected groups in 2020 [2,5,6]. According to the same reports, HIV infection was responsible for around 680,000 deaths worldwide in 2020, compared with 1.3 million in 2010 and 1.9 million in 2004, with a significant decrease (53%) in women and girls [2,6]. However, despite the decrease in HIV incidence and associated mortality worldwide with increased access to potent antiretroviral therapy, incidence rates can nonetheless be alarming in post- or armed conflict countries where sexual violence is used as weapon of war. In 2020, the WHO estimated that 30% of women have been raped, representing a vulnerable group that may be at high risk of HIV infection, particularly adolescent girls, and young women [6]. Sexual violence has become endemic in the eastern DRC, particularly in South Kivu [7]. From September 1999 to March 2021, the teams from the Panzi Hospital have taken care of over 68,774 women survivors of sexual violence (WSSV), among whom only 12% were admitted in less than 72 h and 6% with pregnancies resulting from rape (Panzi Foundation Report 2021).

The South Kivu province, where this study was undertaken, is rife with frequent sexual violence perpetrated on women due to recurrent armed conflicts, especially in rural areas [7]. Furthermore, sporadic cases of sexual violence are reported in the city of Bukavu [8]. These situations may increase sexual transmission of HBV and HIV in this area due to their transmission mode. However, few studies have been conducted on the prevalence of HBV and HIV infections or the burden of HIV/HBV co-infection in South Kivu. This study aims to determine the prevalence of and the factors associated with HBV and HIV infections among WSSV enrolled at one-stop centers in South Kivu. This information is important for predicting the risk of HBV/HIV co-infection and for setting up suitable management of the infections.

## Methods

### Study design, setting, and population

This is a cross-sectional study conducted from May 2018 to May 2020. Participants were recruited from urban and rural one-stop centers of the Panzi Foundation, including the Panzi General Referral Hospital, the Mulamba Hospital Center, and the Bulenga Hospital Center. The Panzi Hospital is the reference center for the health sector in the city of Bukavu. This hospital specializes in several medical areas and today, it is a treatment center for victims and survivors of sexual violence. The hospital receives 5 to 7 female victims of sexual violence every day through the Panzi Foundation, within which the “one-stop center” model of care was developed by Prof [8]. Denis Mukwege, winner of the 2018 Nobel Peace Prize. This model allows victims and survivors of sexual violence to receive holistic care (medical, psychosocial, legal, and socio-economic reintegration) and other services to restore their healthy life needs [8]. Panzi Hospital has partnered up with other one-stop centers, such as the Mulamba Hospital Center, located at about 70 km southwest of Bukavu in the rural area of Walungu, and the Bulenga Hospital Center, located near Goma, 165 km from Bukavu.

### Eligibility criteria

The study population included all women aged 18 years and over with a history of sexual violence of more than 3 months, who received treatment and medical care at one of the above-mentioned one-stop centers. They were admitted for management of various complications related to sexual violence including medical, surgical, gynecological, and social care. A total of 1002 women participants were included in this study. The sample size was determined based on HBV prevalence (5.0%) in the DRC [9], with a precision of ±3% at a 95% confidence level.

### Specimen and data collection

At each one-stop center, prior to administering a questionnaire, an information session was offered to WSSV providing clear explanations, in the local language where necessary, using readily understandable terms about HBV and HIV-related diseases. An informed-consent form was then signed by each participant and the collected information remained anonymous. Sociodemographic and clinical variables were recorded. Clinical variables mainly involved patient history of jaundice, vaccination, transfusion, scarification, tattoo, traditional excision, number of sex partners before and after sexual violence, number of aggressors, and the occurrence of pregnancy; sociodemographic variables included age, origin, tribe, level of schooling, religion, marital status, and occupation. Approximately 10 mL of venous blood was collected in an EDTA tube for rapid screening HIV and HBV tests and results were reported to participants immediately after screening. In addition, dried blood spots (DBS) were prepared in the Medical Research Laboratory (MRL) at the Université Evangélique en Afrique (UEA) in Bukavu by spotting a drop of whole blood on Whatman filter paper. This sample preparation was followed by drying and sealing the DBS in plastic bags for storage at room temperature in presence of a desiccant until transferred to the Viral Hepatitis Laboratory at the Institut Pasteur de Bangui in the Central African Republic (CAR) for further analysis.

### Serological assays

The HBV markers were detected using an enzyme-linked immunosorbent assay (ELISA) provided in DiaSource kits (DiaSource ImmunoAssays S.A., Belgium) according to the manufacturer’s instructions. All samples were tested for the hepatitis B surface antigen (HBsAg), and hepatitis B core antibody (anti-HBc). A confirmatory ELISA test of HBsAg was used for HBsAg-positive samples. HBsAg-negative samples were screened for the HBs antibody (anti-HBs Ab) and HBsAg-positive samples were screened for the hepatitis B e antigen/antibody (HBeAg/anti-HBe Ab) and anti-HBc IgM antibodies. The Murex ELISA kit, (DiaSorin S.p.A., Italy) was used for quality control and confirmation of all positive samples. The presence of HIV was detected using the HIV Ab/Ag Combo ELISA kit (DiaSource ImmunoAssays S.A.) according to the manufacturer’s instructions. After serological testing, all HBV- and HIV-positive samples were transported to the Panzi Hospital laboratory for viral load quantification. This viral load detected HBV DNA and HIV RNA in the blood of patients using Cepheid GeneXpert kits Xpert® HBV Viral Load and Xpert® HIV-1 Qual for HBV and HIV, respectively by following the manufacturer’s recommendations.

### Statistical analysis

Data were collected from a structured survey questionnaire, compiled into a Microsoft Excel 2016 spreadsheet, and then imported into Stata SE 14.0 (Stata Corp LP, College Station, Texas, USA) for clean-up and analysis.

To describe the data, we computed means and their standard deviations (SDs) and medians with interquartile ranges (IQRs) for continuous variables, as appropriate. Categorical variables were summarized as frequencies and their percentages. To compare two means or medians, we used Student’s *t*-test. To compare proportions, we used the Pearson chi-square test or the Fisher exact test for proportions less than or equal to 5. We constructed univariate and multivariate logistic regression models to assess factors associated with HIV-positive and HBV-positive status among WSSV. Variables with a *p*-value 0.2 or lower were included in the model.

The adjusted odds-ratios (AOR) and their 95% confidence intervals (CI) were derived to measure the strength of the association between the variables. All *p*-values were two-sided, and *p*-values less than 0.05 indicated statistical significance.

### Ethical approval and consent to participate

This study obtained approval from the ethics committee of the Catholic University of Bukavu (UCB/CIE/NC/008/2016) reviewed in 2019 by the national health ethics committee (CNES001/DPSK/124PP/2019) and from the ethics and scientific committees of the University de Bangui (22/UB/FACSS/CSVPR/19). Written informed consent was signed by the participant in the interest of knowing her serological status to consider early treatment in case of a positive result, as well as for her participation in the study. The preliminary results for HBV and HIV screening were given to participants immediately after screening and 3 weeks later for definitive result. HIV- and HBV-positive cases were referred to the Panzi Hospital for medical care.

## Results

### Sociodemographic and seroprevalence of HBV and HIV infection among survivors of sexual violence

The average age of the study participants was 37.85 ± 15.11 years and most were 35 years old and older (53.9%). The majority were married (47.7%), Protestant (53.5%) and from rural areas (92.4%) (table 1). Of the 1002 participants, the prevalence rates of HBV markers were 8.9% (95% CI: [7.2 – 10.8%]), 32.1% [29.3–35.0%], and 14.5% [12.3–16.8%] for HBsAg, anti-HBc and anti-HBs antibodies, respectively. Of the 89 HBsAg-positive participants, 17 (19.1%) were also HBeAg-positive.

**Table 1.**
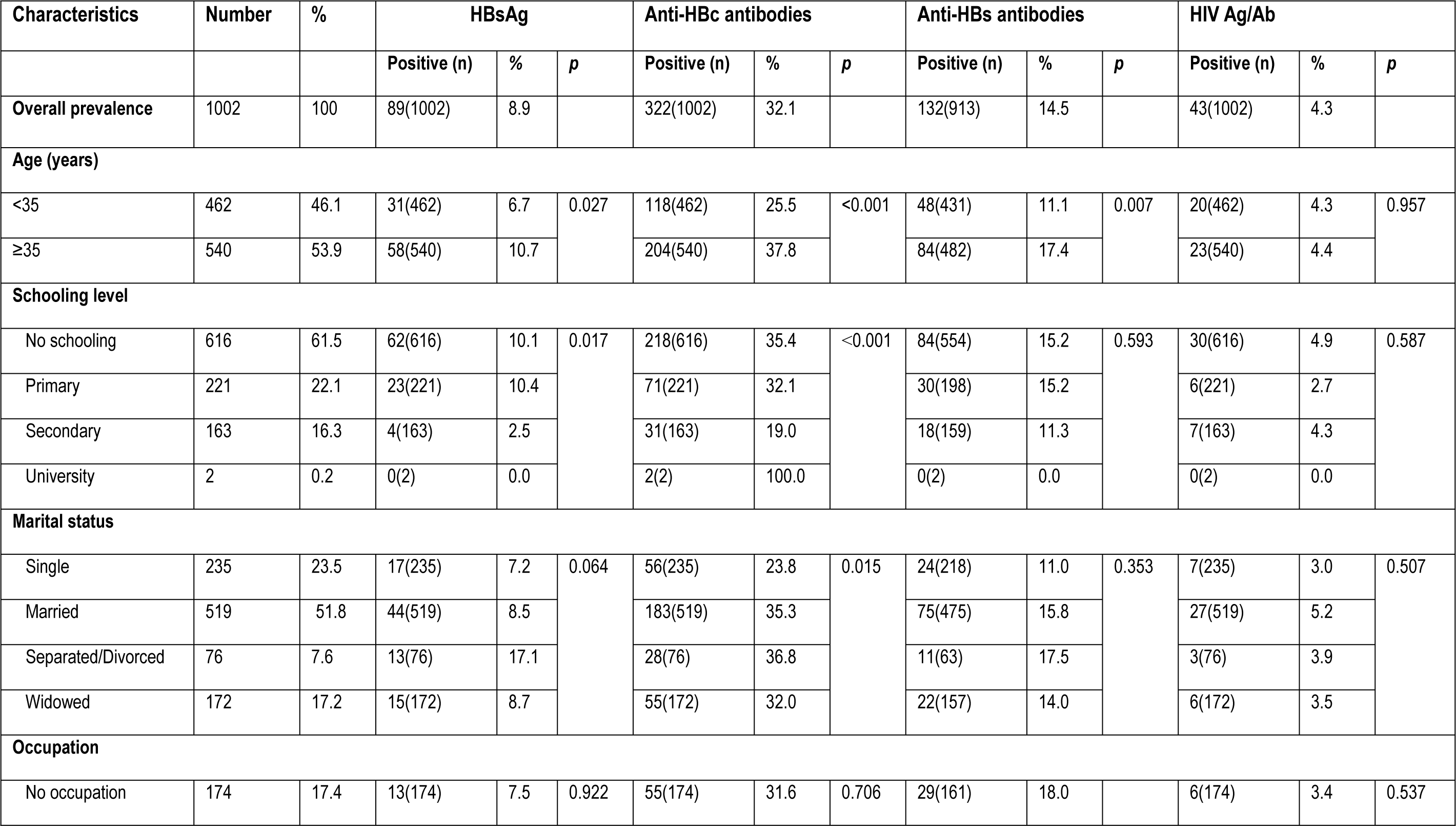
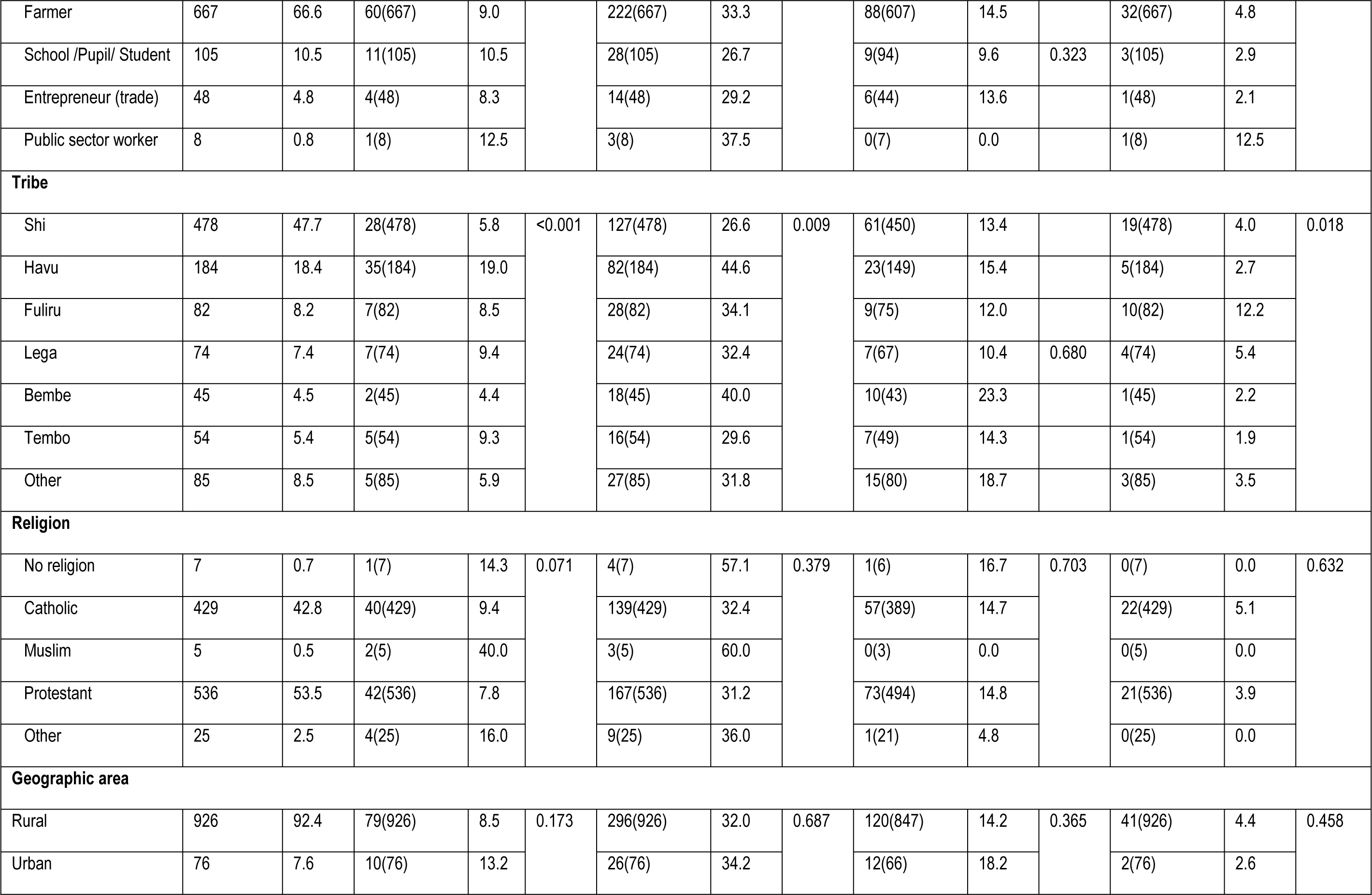
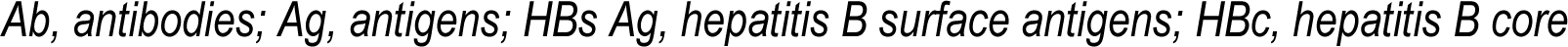
Sociodemographic characteristics and prevalence of hepatitis B virus (HBV) and human immunodeficiency virus (HIV) among survivors of sexual violence in South Kivu.

The mean age was 40.57±14.99 years in the HB-positive group, statistically significantly higher than the overall mean age (*p*=0.025). Furthermore, participants aged 35 years and older were more frequently HBsAg-positive than those under 35 years (*p*=0.027). A statistical difference was observed in participants with no formal schooling or only or primary-school education (10.1 and 10.4%, respectively; *p*=0.017) compared with those with secondary- or university-level education. However, there was no statistical difference in the prevalence of HBV infection according to marital status or occupation, despite the slightly higher number of infection cases among separated/divorced women and public sector workers (17.1 and 12.5%; *p*=0.064 and *p*=0.922, respectively). Participants from the Havu tribe showed higher prevalence compared with participants from other tribes (19.1%; *p*<0.001). Religion and geographic origin of participants were not statistically associated with HBsAg positivity.

Similarly, for the hepatitis B contact marker (anti-HBc), the 35+ age group had a high prevalence compared with the under-35 age group (37.8% vs. 25.5%; *p*<0.001). For anti-HBs antibodies, a statistical difference was also observed between the age groups, with higher immunization in the over-35 age group (17.4% vs. 11.1%; *p*=0.007) compared with those under 35 years of age.

A total of 43 of the 1002 participants (4.3% [3.1%–5.7%]) tested positive for HIV. The Fuliru tribe was more frequently associated with HIV infection than the other tribes (12.2%; *p*=0.018). Although HIV infection appeared high, it was not statistically significant among women aged 35 years and over (4.4%), with no schooling (4.9%), married (5.2%), public sector workers (12.5%), Catholic (5.1%), or from rural areas (4.4%) (Table 1). Regarding HBV-HIV co-infection, the prevalence was 0.5%.

### Clinical history, and HBV and HIV infections among survivors of sexual violence

For HBV infection, a statistically significant association was found between the presence of HBsAg and multiple aggressors (11.0%; *p*=0.022). The absence of HBV vaccination was statistically associated with the presence of HBsAg (*p*=0.011). There was no statistical difference between the prevalence of HBV infection and other factors. However, participants with a history of tattoo, traditional genital excision, multiple sexual partners before sexual violence and current multiple sexual partners had slightly higher prevalences of HBV (10.3%, 10.6%, 10.0%, and 9.1%, respectively) (Table 2). For HIV infection, no association was found with those factors.

**Table 2.** Clinical history and hepatitis B virus (HBV) and human immunodeficiency virus (HIV) infections among women survivors of sexual violence.

| Characteristics | Number | HBsAg |  |  | HIV Ag/Ab |  |  |
| --- | --- | --- | --- | --- | --- | --- | --- |
|  | N(%) | Positive n(%) | Negative n(%) | *p* | Positive n(%) | Negative n(%) | *p* |
| **Pregnancy** |  |  |  |  |  |  |  |
| Yes | 61(6.1) | 5(8.2) | 56(91.8) | 0.846 | 2(3.3) | 59(96.7) | 0.687 |
| No | 941(93.9) | 84(8.9) | 857(91.1) |  | 41(4.4) | 900(95.6) |  |
| **History of jaundice** |  |  |  |  |  |  |  |
| Yes | 23(2.3) | 2(8.7) | 21(91.3) | 0.975 | 1(4.3) | 22(95.7) | 0.989 |
| No | 979(97.7) | 87(8.9) | 892(91.1) |  | 42(4.3) | 937(95.7) |  |
| **Vaccination** |  |  |  |  |  |  |  |
| Yes | 6(0.6) | 0(0.0) | 6(100.0) | 0.011 | 0(0.0) | 6(100.0) | 0.603 |
| No | 996(99.4) | 89(8.9) | 907(91.1) |  | 43(4.3) | 953(95.7) |  |
| **Transfusion** |  |  |  |  |  |  |  |
| Yes | 47(4.7) | 1(2.1) | 46(97.9) | 0.095 | 1(2.1) | 46(97.9) | 0.453 |
| No | 955(95.3) | 88(9.2) | 867(90.8) |  | 42(4.4) | 913(95.6) |  |
| **Condom use** |  |  |  |  |  |  |  |
| Never | 974(97.2) | 88(9.0) | 886(91.0) | 0.316 | 41(4.2) | 933(95.8) | 0.450 |
| Yes | 28(2.0) | 1(3.5) | 27(96.5) |  | 2(7.1) | 26(92.9) |  |
| **Tattoo** |  |  |  |  |  |  |  |
| Yes | 39(3.9) | 4(10.3) | 35(89.7) | 0.758 | 2(5.1) | 37(94.9) | 0.793 |
| No | 963(96.1) | 85(8.8) | 878(91.2) |  | 41(4.3) | 922(95.7) |  |
| Scarification |  |  |  |  |  |  |  |
| Yes | 28(2.8) | 2 (7.1) | 26(92.9) | 0.743 | 0(0.0) | 28(100.0) | 0.256 |
| No | 974(97.2) | 87(8.9) | 887(91.1) |  | 43(4.4) | 931(95.6) |  |
| Traditional excision |  |  |  |  |  |  |  |
| Yes | 123(12.3) | 13(10.6) | 110(89.4) | 0.483 | 4(3.3) | 119(96.7) | 0.544 |
| No | 879(87.7) | 76(8.6) | 803(91.4) |  | 39(4.4) | 840(95.6) |  |
| Multiple sex partners before sexual violence |  |  |  |  |  |  |  |
| Yes | 609(60.8) | 61(10.0) | 548(90.0) | 0.174 | 28(4.6) | 581(95.4) | 0.633 |
| No | 393(39.2) | 28(7.1) | 365(92.9) |  | 15(3.8) | 378(96.2) |  |
| Current multiple sex partners |  |  |  |  |  |  |  |
| Yes | 536(53.5) | 49(9.1) | 487(90.9) | 0.824 | 20(3.7) | 516(96.3) | 0.354 |
| No | 466(46.5) | 40(8.6) | 426(91.4) |  | 23(4.9) | 443(95.1) |  |
| Number of aggressors |  |  |  |  |  |  |  |
| Single | 521(52.0) | 36(6.9) | 428(93.1) | 0.022 | 21(4.0) | 499(96.0) | 0.672 |
| Multiple | 481(48.0) | 53(11.0) | 428(89.0) |  | 22(4.6) | 459(95.4) |  |

Ab, antibodies; Ag, antigens; HBs Ag, hepatitis B surface antigens; HBc, hepatitis B core; virus; HIV, human immunodeficiency virus.

### Risk factors and HBV infection

The results showed that HBs-positive status was associated with women aged 35 or older (AOR=1.83 [1.02–3.32]; *p*=0.041), having no schooling (AOR=4.14 [1.35–12.62]; *p*=0.012) or primary schooling only (AOR=4.88 [1.61–14.75]; *p*=0.005), being part of the Havu tribe (AOR=4. 84 [1.08–21.64]; *p*=0.039), and multiple aggressors (AOR=1.76 [1.09–2.84]; *p*=0.019) (Table 3).

**Table 3.** Univariate and multivariate analysis of factors associated with hepatitis B status among survivors of sexual violence.

| Characteristics | COR [95% CI] | p-Value | AOR [95% CI] | p-value |
| --- | --- | --- | --- | --- |
| **Age group (years)** |  |  |  |  |
| <35 | 1 (reference) |  | 1 (reference) |  |
| ≥35 | 1.67 [1.06–2.63] | 0.027 | 1.83 [1.02–3.32] | 0.041 |
| **Marital status** |  |  |  |  |
| Single | 1 [reference] |  | 1 [reference] |  |
| Married/Homemaker | 1.18 [0.66–2.12] | 0.562 | 0.93 [0.47–1.85] | 0.848 |
| Separated/Divorced | 2.64 [1.21–5.74] | 0.014 | 2.15 [0.84–5.47] | 0.107 |
| Widowed | 1.22 [0.59–2.52] | 0.582 | 0.74 [0.28–1.96] | 0.547 |
| **Schooling level** |  |  |  |  |
| None | 4.44 [1.59–12.41] | 0.004 | 4.14 [1.35–12.62] | 0.012 |
| Primary | 4.61 [1.56–13.62] | 0.006 | 4.88 [1.61–14.75] | 0.005 |
| Secondary | - | - | - | - |
| University | 1 (reference) |  | 1 (reference) |  |
| **Religion** |  |  |  |  |
| None | 1 (reference) |  |  |  |
| Catholic | 0.61 [0.07–5.25] | 0.659 | – | – |
| Muslim | 4.00 [0.25–63.95] | 0.327 | – | – |
| Protestant | 0.51 [0.59–4.33] | 0.538 | – | – |
| Other | 1.14 [0.10–12.24] | 0.912 | – | – |
| **Blood transfusion** |  |  |  |  |
| No | 1 (reference) |  | 1 (reference) |  |
| Yes | 0.21 [0.29–1.57] | 0.130 | 0.19 [0.02–1.45] | 0.110 |
| **Aggressors** |  |  |  |  |
| Single | 1 (reference) |  | 1 (reference) |  |
| Multiple | 1.66 [1.07–2.59] | 0.023 | 1.76 [1.09–2.84] | 0.019 |
| **Tribe** |  |  |  |  |
| Bembe | 1 (reference) |  | 1 (reference) |  |
| Fuliru | 2 [0.39–10.09] | 0.398 | 2.25 [0.43–11.70] | 0.334 |
| Havu | 5.05 [1.16–21.85] | 0.030 | 4.84 [1.08–21.64] | 0.039 |
| Lega | 2.24 [0.44–11.32] | 0.327 | 2.68 [0.51–14.03] | 0.241 |
| Shi | 1.33 [0.30–5.80] | 0.698 | 1.42 [0.31–6.34] | 0.642 |
| Tembo | 2.19 [0.40–11.89] | 0.362 | 2.89 [0.51–16.32] | 0.228 |
| Other | 1.34 [0.25–7.21] | 0.731 | 1.56 [0.28–8.72] | 0.609 |
| **Use of condoms** |  |  |  |  |
| Yes | 1 (reference) |  | 1 (reference) |  |
| No | 2.68 [0.36–19.97] | 0.336 | 1.84 [0.22–14.75] | 0.565 |

\*COR: Crude odds ratio; \*\*AOR: Adjusted odds ratio. In this table, we present the crude and adjusted odds ratios.

For HIV, after adjustment of the logistic regression model, we did not find any statistically significant associations between the risk factors studied and HIV status among WSSV (*p*>0.05).

### Level of HBV and HIV viral loads in positive participants

Out of a total of 89 HBsAg-positive samples, the mean viral load was 4949423.89 ± 22250009 IU/mL; 38.2% of the samples had an undetectable viral load, 39.3% a low viral load (˂2000 IU/mL), and 22.5% a high viral load (≥2000 IU/mL). For HIV, for those participants who tested positive for the first time, the average viral load was 227924.24 ± 734065.81 copies/mL (table 4. The distribution of the HIV viral load showed a low (˂1000 copies/mL) viral load for 35.2% of the patients and a high (≥1000 copies/ml) for 64.8% (Table 4).

**Table 4.** HBV and HIV viral load results in participants who tested positive.

| Virological parameters | Number | % |
| --- | --- | --- |
| **HBV DNA viral load (UI/mL)** | **N=89** |  |
| Mean±SD | 4949423.89 ± 22250009 |  |
| Undetectable | 34 | 38.2 |
| < 2000 | 35 | 39.3 |
| ≥ 2000 | 20 | 22.5 |
| **HIV RNA viral load (copies/mL)** | **N=34** |  |
| Mean±SD | 227924.24 ± 734065.81 |  |
| < 1000 | 12 | 35.2 |
| ≥ 1000 | 22 | 64.8 |

## Discussion

In this study, the overall prevalence of HBsAg was 8.9% [7.2%-10.8%] and 32.1% [29.3%–35.0%] for anti-HBc antibody. These results demonstrate the high burden of HBV in this vulnerable population. The prevalence of HBV infection varies across regions and different types of population in the DRC [9]. Previous studies on HBV infection have focused on blood donors in DRC different regions, reporting HBsAg prevalence rates ranging from 6.8% to 9.2%, but in other population groups, prevalence seems to be much lower [10–17]. For example, one study in the Maniema province reported a prevalence of 5.9% for HBsAg in pregnant women [18]. But the results of 2013-2014 Demographic and Health Survey on the general population showed a prevalence of 3.3% in adults and 2.2% in children [9]. In contrast, an HBsAg prevalence of 24.6% was reported on samples from 2003 to 2012 in the yellow fever surveillance program from patients with acute febrile jaundice and suspected yellow fever [19].

The difference in hepatitis B prevalence rates in these different regions of the DRC can likely be attributed to the exposure to different risk factors according to population type, and probably arises from the absence of a viral hepatitis control and prevention program, as well as the fact that hepatitis B vaccination is limited to infants only. This variation in prevalence is also found in sub-Saharan Africa, where the rate of HBV infection, which is often very high, also varies with the country, the studied populations, and the used techniques [20–37]. For example, the overall prevalence of 42.3% for anti-HBc antibody with 15.5% for HBsAg in students with high sexual activity in Bangui [23].

Our investigation also revealed that 14.5% of the study population shows anti-HBs antibodies. Of the participants with this immunization marker, only 11.4% had a history of hepatitis B vaccination when tested within 72 h of sexual violence, and 88.6% had no history of previous hepatitis B vaccination. In the latter, the total anti-HBc antibody was present, clearly demonstrating that they had been in contact with the virus itself. An association was found between the rate of immunization against HBV infection and the over-35 years age group. Therefore, it is difficult to demonstrate precisely when the infection was acquired with respect to sexual abuse but given the high proportion of antibody detection in older patients, the hypothesis of infection acquired in conjunction with sexual abuse is highly plausible. Unfortunately, the antibody titer was not determined in this study. The anti-HBs antibody titer for HBV has been shown to decrease with age according to the age of the infection. Thus, the proportion of older individuals with anti-HBs antibodies at ≥10 IU/mL may be low if the infection was acquired in childhood, although they may nevertheless remain immunized against the infection [38].

For HIV infection, a prevalence of 4.3% [3.1%–5.7%] was found using fourth-generation ELISA tests. This prevalence is very high for this population category compared to that reported in a previous study among blood donors in Bukavu, which showed a low prevalence using rapid tests [11]. However, before the introduction of blood safety protocols in Kisangani, a prevalence of 4.7% was reported in this same group [15]. Nevertheless, our study demonstrates, the high prevalence of HIV among sexually abused patients in South Kivu province.

The link between sexual violence against women and girls and HIV acquisition has been shown in several studies [39]. For example, in Uganda, 15–49 year-old women who had experienced sexual violence showed a 1.55 risk of HIV infection compared with those who had never experienced violence [40]. Similarly, in a study in South Africa, young women aged 15–24 years who reported multiple episodes of sexual violence were 1.51 more at risk to acquire HIV than women with one or no episodes of sexual violence; furthermore, an estimated 12% of new HIV infections could be attributed to sexual violence [41].

The same findings were reported in a cross-sectional study of 28,000 married women aged 15–49 years in India, with a 4-fold higher risk of being HIV-positive among those who had experienced physical and sexual violence by their partners than those who had not [42]. Although studies of HIV among women who have experienced sexual violence are rare in the DRC, HIV prevalence is often very high (7.5%) among female sex workers. This high prevalence contrasts with the overall HIV incidence in the general population, which is estimated to have decreased from 2010 to 2020 from 1.2% to 0.7%, with a national prevalence of 1.1% among women aged 15–49 years [43]. Furthermore, prevalence may be underestimated considering the low rate of HIV testing in different groups and key populations, the use of rapid tests in routine screening, and diagnoses that sometimes produce false negatives with a risk of undetected HIV cases [44]. It has been well documented that women are two to four times more likely than men to contract the virus through unprotected vaginal penetration [45–47]. Sexual abuse of women and girls by one or more individuals is a direct risk factor for HIV transmission. The risk factors associated with HIV infection observed in this study were as follows: age less than or equal to 25 years, belonging to the Fulero tribe, occasional condom use, and number of assailants greater than or equal to 5. Women with a history of sexual assaults with more than 5 men in their lifetime had a higher frequency of HIV infection than those with fewer than 5 abusers. These results are consistent with studies that have shown that HIV is more prevalent among young women, sex workers, those with multiple sexual partners, those who have experienced physical or sexual violence, and those with low levels of education, but other factors such as poverty and early marriage are also implicated [47–50].

The main sources of HIV transmission vary by country. Risks found in sexual behavior include age at first intercourse [51], being sexually active, having multiple sexual partners, having unprotected sex, having unprotected sex with strangers [52], having sex during or after alcohol use [53], sexual violence [45], and transactional sex [54].

Addressing violence against women and gender equality has been identified by UNAIDS in the AIDS Investment Framework as essential for an effective response to the HIV epidemic [55]. Addressing gender-based violence (GBV) could have a significant impact on the HIV epidemic. Indeed, the national HIV program needs to recognize the following aspects: the direct and indirect implications of GBV in the mechanism of HIV dissemination, the importance of perpetrator dynamics, and that GBV reduction should be part of HIV prevention programs. Effective interventions are likely to include a structural component and a sexual violence awareness component.

Our study also revealed the prevalence of HBV/HIV co-infection that was 0.5%. This prevalence appears to be low, because most studies on co-infection in sub-Saharan Africa always report high prevalence rates [56]. Nevertheless, this prevalence varies across regions defined by HBV endemicity. In regions such as sub-Saharan Africa and East Asia, where HBV prevalence is high, most HBV infections occur perinatally or in early childhood through close household contact, medical procedures, or cultural practices such as scarification or tattooing [57]. HBV infections are therefore more likely to progress to chronic infections, resulting in a high prevalence of chronic HBV infection among young people at risk for sexual HIV [58].

HBV and HIV quantifications were performed on samples from our study populations using the automated GeneXpert® system. The HBV viral load results showed that 77.5% had undetectable or low viral load and 22.5% had a high viral load (≥2000 IU/mL). The 77.5% for whom the viral load was undetectable or low were not likely to be put on antiviral therapy. However, the 22.5% with high viral load should be treated therapeutically for infection, following further testing. This quantification test must be made available and accessible to all patients in South Kivu to avoid shortages of antivirals or high expenses related to the routine therapeutic management of anyone infected with HBV. Indeed, it is known that more than 90% of infected adults spontaneously recover from acute viral hepatitis B, with the disappearance of HBsAg after about 6 months [59,60]. Accurate measurement of HBV DNA levels in the blood is essential to diagnose HBV infection, establish the prognosis of HBV-related liver disease, and guide therapeutic decision-making to determine whether to initiate antiviral therapy and monitor the virological response to antiviral therapy and emergence of resistance [61,62].

In resource-limited countries such as the DRC, the ability to determine the HBV viral load is a major issue for the therapeutic management of patients, because the technical facilities recommended by international guidelines are not available and/or accessible [63]. Quantifying HBV DNA in the blood is one of the essential tests to establish the prognosis of infection and support any therapeutic decisions [61,62].

For the 34 subjects in whom the HIV viral load was determined, 64.8% had a high viral load (≥1000 copies/mL) and 35.2% a low viral load (<1000 copies/mL). These viral loads show that HIV testing is still a real problem in this area of the world due to the inaccessibility of ELISA tests, which clearly have added value for the detection and diagnosis of HIV cases. The dearth of modern technical facilities contributes to the underestimation of HIV cases following the use of rapid tests. In this study, the positive cases had a detectable HIV viral load, except for nine cases that were not tested due to the insufficient amount of blood sample.

Our results clearly show the importance of using comprehensive tests for diagnosis of HBV and HIV in a vulnerable population group. However, not all tests to establish a complete diagnosis of liver diseases, such as transaminase and other complementary tests, were carried out, which thus constitutes a limitation of this study. Determining viral load remains an essential tool for the management of HBV infection.

## Conclusion

This study shows a high prevalence of HBV and HIV infections among girls and women who have survived sexual violence in South Kivu, despite the low national prevalence of HIV and the intermediate endemicity of HBV infection in the DRC. In this study, the ELISA technique revealed that the prevalence of these infections is underestimated in South Kivu where girls and women have been exposed to sexual violence for many years due to repetitive wars. We suggest that actions to combat sexual violence against girls and women be integrated into strategies to combat HIV/AIDS and viral hepatitis B.

## Contributors

JBN and NPJK supervised the study. PBB, JBN, and NPJK designed the study, wrote the first manuscript. PBB, TKM, CMM, and BMM contributed to the laboratory analyses. PBB and OM performed the statistical analyses. PBB, DBM, AKB, GKB and CMM contributed to the collection of clinical data and the medical management of patients. PBB, JNB, NPJK, JPM, DMM contributed to the critical revision of the manuscript. All authors contributed to the interpretation of the results, approved the final manuscript and accept responsibility for the decision to submit it for publication.

## Data Sharing

All data were encoded and stored in an electronic database. Follow-up sheets were securely stored at the ICART/Panzi Foundation Research Center and accessible only to the principal investigator of the study. These data include epidemiological, clinical and therapeutic data. As these data are anonymized for the protection of participants according to the health ethics guidelines of our region, we cannot make them available to third parties.

## Data Availability

All data were encoded and stored in an electronic database. Follow-up sheets were securely stored at the ICART/Panzi Foundation Research Center and accessible only to the principal investigator of the study, Dr Parvine Basimane Bisimwa. These data include epidemiological, clinical and therapeutic data. As these data are anonymized for the protection of participants according to the health ethics guidelines of our region, we cannot make them available to third parties. Nevertheless, all these data are available from Parvine Bisimane Bisimwa at

## Acknowledgements

The research presented in this publication was funded by the World Bank through the Panzi Foundation. We would like to thank the viral hepatitis laboratory team at the Institut Pasteur de Bangui for all the support received in conducting the laboratory analyses and facilitating the technical operations. This work is a research collaboration between the Panzi Foundation and the Institut Pasteur de Bangui. We are very grateful for the involvement of the director of the three ’One Stop Center’ hospital. We would also like to thank Mr Giscard Wilfried Koyaweda, Mr Rodrigue Balthazar Basengere Ayagirwe, Mr Ahadi Bwihangane Birindwa, Mr Patrick Ntagereka Bisimwa for their contribution in the re-reading of this manuscript.

